# Effectiveness of CoronaVac among healthcare workers in the setting of high SARS-CoV-2 Gamma variant transmission in Manaus, Brazil: A test-negative case-control study

**DOI:** 10.1101/2021.04.07.21255081

**Authors:** Matt D.T. Hitchings, Otavio T. Ranzani, Mario Sergio Scaramuzzini Torres, Silvano Barbosa de Oliveira, Maria Almiron, Rodrigo Said, Ryan Borg, Wade L. Schulz, Roberto Dias de Oliveira, Patricia Vieira da Silva, Daniel Barros de Castro, Vanderson de Souza Sampaio, Bernardino Cláudio de Albuquerque, Tatyana Costa Amorim Ramos, Shadia Hussami Hauache Fraxe, Cristiano Fernandes da Costa, Felipe Gomes Naveca, Andre M. Siqueira, Wildo Navegantes de Araújo, Jason R. Andrews, Derek A.T. Cummings, Albert I. Ko, Julio Croda

## Abstract

**Background:** Severe acute respiratory syndrome coronavirus 2 (SARS-CoV-2) variant, Gamma, emerged in the city of Manaus in late 2020 during a large resurgence of coronavirus disease (COVID-19), and has spread throughout Brazil. The effectiveness of vaccines in settings with widespread Gamma variant transmission has not been reported.

**Methods:** We performed a matched test-negative case-control study to estimate the effectiveness of an inactivated vaccine, CoronaVac, in healthcare workers (HCWs) in Manaus, where the Gamma variant accounted for 86% of genotyped SARS-CoV-2 samples at the peak of its epidemic. We performed an early analysis of effectiveness following administration of at least one vaccine dose and an analysis of effectiveness of the two-dose schedule. The primary outcome was symptomatic SARS-CoV-2 infection.

**Findings:** For the early at-least-one-dose and two-dose analyses the study population was, respectively, 53,176 and 53,153 HCWs residing in Manaus and aged 18 years or older, with complete information on age, residence, and vaccination status. Among 53,153 HCWs eligible for the two-dose analysis, 47,170 (89%) received at least one dose of CoronaVac and 2,656 individuals (5%) underwent RT-PCR testing from 19 January, 2021 to 13 April, 2021. Of 3,195 RT-PCR tests, 885 (28%) were positive. 393 and 418 case-control pairs were selected for the early and two-dose analyses, respectively, matched on calendar time, age, and neighbourhood. Among those who had received both vaccine doses before the RT-PCR sample collection date, the average time from second dose to sample collection date was 14 days (IQR 7-24). In the early analysis, vaccination with at least one dose was associated with a 0.50-fold reduction (adjusted vaccine effectiveness (VE), 49.6%, 95% CI 11.3 to 71.4) in the odds of symptomatic SARS-CoV-2 infection during the period 14 days or more after receiving the first dose. However, we estimated low effectiveness (adjusted VE 36.8%, 95% CI -54.9 to 74.2) of the two-dose schedule against symptomatic SARS-CoV-2 infection during the period 14 days or more after receiving the second dose. A finding that vaccinated individuals were much more likely to be infected than unvaccinated individuals in the period 0-13 days after first dose (aOR 2.11, 95% CI 1.36-3.27) suggests that unmeasured confounding led to downward bias in the vaccine effectiveness estimate.

**Interpretation:** Evidence from this test-negative study of the effectiveness of CoronaVac was mixed, and likely affected by bias in this setting. Administration of at least one vaccine dose showed effectiveness against symptomatic SARS-CoV-2 infection in the setting of epidemic Gamma variant transmission. However, the low estimated effectiveness of the two-dose schedule underscores the need to maintain non-pharmaceutical interventions while vaccination campaigns with CoronaVac are being implemented.

**Funding:** Fundação Oswaldo Cruz (Fiocruz); Municipal Health Secretary of Manaus

**Research in Context:** *Evidence before this study:* We searched PubMed for articles published from inception of the pandemic until April 3, 2021, with no language restrictions, using the search terms “P.1” AND “vaccine” AND “SARS-CoV-2”. Additionally, we searched for “CoronaVac” AND “SARS-CoV-2”. Early studies have found plasma from convalescent COVID-19 patients and sera from vaccinated individuals have reduced neutralisation of the SARS-CoV-2 variant, Gamma or P.1, compared with strains isolated earlier in the pandemic. Pfizer BNT162b2 mRNA, Oxford-AstraZeneca ChAdOx1, and CoronaVac are the only vaccines for which such data has been published to date. No studies reported effectiveness of any vaccine on reducing the risk of infection or disease among individuals exposed to P.1 or in settings of high P.1 transmission.

*Added value of this study:* This study finds that vaccination with CoronaVac was 49.4% (95% CI 13.2 to 71.9) effective at preventing COVID-19 in a setting with likely high prevalence of the Gamma Variant of Concern. However, an analysis of effectiveness by dose was underpowered and failed to find significant effectiveness of the two-dose schedule of CoronaVac (estimated VE 37.1%, 95% CI -53.3 to 74.2).

*Implications of all the available evidence:* These findings are suggestive for the effectiveness of CoronaVac in healthcare workers in the setting of widespread P.1 transmission but must be strengthened by observational studies in other settings and populations. Based on this evidence, there is a need to implement sustained non-pharmaceutical interventions even as vaccination campaigns continue.

## Introduction

The P.1, or Gamma, variant of severe acute respiratory syndrome coronavirus 2 (SARS-CoV-2) emerged in Manaus, Brazil, in November 2020^1–3^ and has since spread globally to 52 countries as of 8 June, 2021.^4^ The World Health Organisation declared the Gamma variant as a *Variant of Concern*^5^ given the evidence for its increased transmissibility^3^ and mutations shared with other variants of concern. Brazil has recently experienced a COVID-19 resurgence, during which 6,453,057 cases and 151,467 deaths were reported between 1 December 2020 and 31 March 2021.^6^ A critical question is whether available vaccines in Brazil and South America are effective against COVID-19 in the context of Gamma variant transmission.

Concerns have been raised that available vaccines have reduced immunogenicity against the Gamma variant. The variant has three mutations, K417N, E484K and N501Y, in the ACE2 binding site of the SARS-CoV-2 S1 protein which have been speculated to promote immune escape.^3^ *In vitro* studies have found decreased sero-neutralisation of the Gamma variant in individuals infected with non-Gamma strains and vaccinated individuals.^7–13^ However, evidence is lacking on whether available vaccines are effective against clinical and infection outcomes associated with the Gamma variant and in settings of Gamma variant transmission in Brazil and beyond.

As part of its vaccination campaign, Brazil has administered CoronaVac, an inactivated vaccine.^14–16^ CoronaVac was found to have an efficacy of 50% and 84% against, respectively, symptomatic COVID-19 and COVID-19 requiring medical assistance in a randomised controlled trial (RCT) in healthcare workers conducted in Brazil prior to the emergence of the Gamma variant^16^ and has been approved by the WHO Emergency Use Listing procedure.^17^ However, the effectiveness of CoronaVac in the real-world setting and in regions of Gamma variant transmission is unknown. We performed a test-negative case-control study^18,19^ on the effectiveness of CoronaVac in healthcare workers (HCWs) from Manaus, which was among the first cities in Brazil to aggressively implement vaccination. Herein, we report early findings on the effectiveness of administering at least one dose of the two-dose schedule, in response to the need to evaluate current vaccination efforts as they are being widely implemented, and a subsequent analysis of effectiveness of the complete two-dose schedule.

## Methods

### Study setting

Manaus is a city with 2.2 million inhabitants in the Amazon Basin of Brazil.^20^ Manaus is an important economic centre in the North of Brazil, with a human development index of 0.737 and high income inequality (Gini index, 0.634).^21^ As of 2 April, 2021, 160,803 cases (cumulative incidence, 7,367 per 100,000 population) and 8,432 COVID-19 associated deaths (cumulative mortality, 386 per 100,000 population) were reported in Manaus during the course of an initial epidemic in March 2020 and a second larger epidemic in late November 2020 (Supplementary Figure 1, Figure 1).^22^ Reported incidence likely represents an underestimate of true incidence due to lack of access to testing, and cumulative incidence in Manaus has been estimated to be significantly higher than reported.^23,24^ The second epidemic was associated with the emergence and spread of the Gamma variant,^1–3^ which accounted for 66% (143 of 247) of SARS-CoV-2 samples genotyped as part of surveillance during the peak of the epidemic in January 2021 (Supplementary Figure 2).^2^ The Municipal Secretary of Health of Manaus (SEMSA) initiated vaccination with CoronaVac and Oxford-AstraZeneca (ChAdOx1) on 19 January 2021; CoronaVac has been used in >97% of the vaccinations of HCWs (Figure 1).

**Figure 1.**
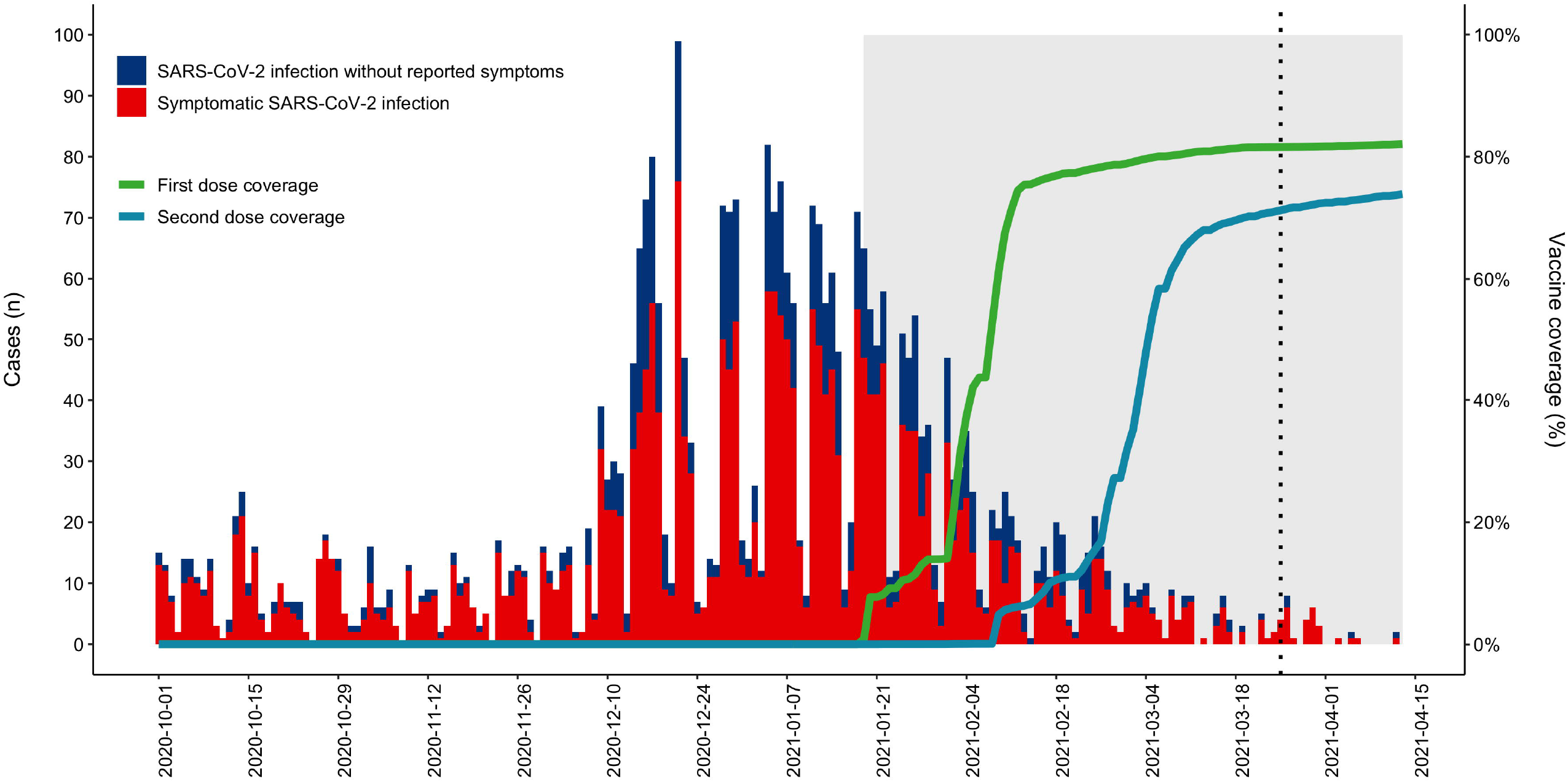
SARS-CoV-2 infections and vaccination coverage amongst 67,718 healthcare workers (HCW) in Manaus, Brazil between 1 October 2020 to 13 April 2021. Daily RT-PCR confirmed SARS-CoV-2 infections with and without COVID-19 symptoms are shown as red and blue bars, respectively. Green and blue lines depict the daily cumulative proportion of the 67,718 HCWs who received respectively, a first and second dose of a COVID-19 vaccine. The grey shade denotes the study period, which began with the initiation of the vaccine campaign on 19 January, 2021 and ended on 25 March, 2021 for the early analysis and on 13 April, 2021 for the two-dose analysis.

HCWs generally had more access to testing than the general population, regardless of symptoms. We evaluated all HCWs in Manaus, including those in hospitals, primary care, general, and specialized units. Testing availability was heterogeneous across settings and location. There was no specific testing or screening policy in place for HCWs, and asymptomatic individuals being tested could arise from contact tracing or voluntary screening. Initially, vaccination of HCWs was stratified by risk of SARS-CoV-2 infection because of the scarcity of vaccines; the priority order was HCWs working in the ICU, ER, COVID-19 wards, and finally administrative staff.

### Study design

We conducted a retrospective, test-negative, matched case-control study to estimate the effectiveness of CoronaVac in reducing the odds of primary and secondary outcomes of, respectively, symptomatic and all RT-PCR-confirmed SARS-CoV-2 infections. The study population was HCWs who had a residential address in Manaus, aged ≥ 18 years on 19 January, 2021, and with complete information, which was consistent between data sources, on age, sex, neighbourhood (*bairro*) of residence, and vaccination status and dates (Figure 2).

**Figure 2.**
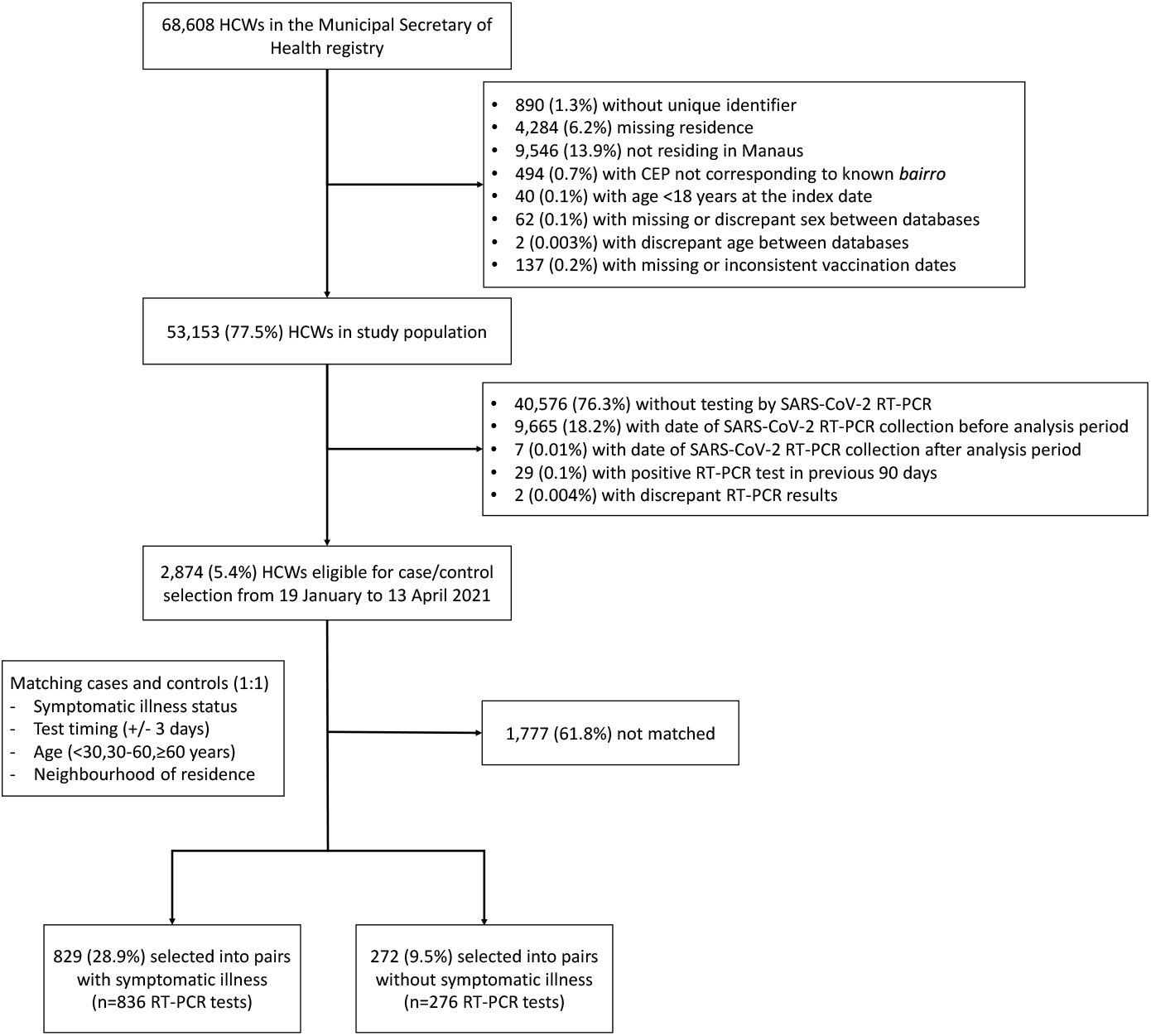
Flowchart for case and control selection.

To ensure timely communication of results with potential public health benefit, we planned an analysis assessing the effectiveness of receiving at least one dose. For this early analysis, we selected cases and matched controls who had a positive and negative SARS-CoV-2 RT-PCR test result, respectively, during the study period of 19 January to 25 March, 2021. In addition, we planned an analysis of effectiveness of the two-dose schedule, for which the study period was extended to 13 April, 2021.

The study design and statistical analysis plan were specified in advance of extracting information from data sources and are described in a publicly available protocol (https://github.com/juliocroda/VebraCOVID-19) and the supplementary file. The study was approved by the Ethical Committee for Research of Federal University of Mato Grosso do Sul (CAAE: 43289221.5.0000.0021).

### Data sources

We identified the study population from the SEMSA registry of employed HCWs in Manaus (Figure 2). For the purpose of extracting information for study population eligibility, case and control section, matching criteria, secondary outcomes and covariates, we integrated data from the following sources: the national laboratory testing registry (“GAL”); the national registry of users of the universal health system(“CadSUS”); the national surveillance system of suspected COVID-19 cases (“e-SUS”); the national surveillance database of severe acute respiratory illnesses (“SIVEP-Gripe”); and the SEMSA COVID-19 vaccination registry. These are surveillance systems for the whole country and notification is compulsory. The early analysis and two-dose analysis were implemented with data that were accessed on 1 April and 20 April, 2021 respectively, and censored after 25 March and 13 April, 2021 to account for reporting delays.

### Selection of cases and matched controls

Cases were selected from the study population who had a SARS-CoV-2 infection, defined as a positive SARS-CoV-2 RT-PCR test result from a respiratory sample that was collected during the study period and the absence of a positive test in the preceding 90-day period. Controls were selected from the study population who did not have a SARS-CoV-2 infection, defined as a negative SARS-CoV-2 test result from a respiratory sample that was collected during the study period and the absence of a positive test in the preceding 90-day period and subsequent 14-day period. For both cases and controls, we excluded PCR tests that were performed after the individual received any dose of ChAdOx1, as this study was limited to assessment of the effectiveness of CoronaVac.

We matched one test-negative control to each case according to symptomatic illness status at time of testing; a time window of ±3 days between the case sample collection date; age category defined as <30, ≥30 and <60, and ≥60 years; and neighbourhood of residence. Symptomatic illness was defined as the presence of one or more reported COVID-19 related symptom^25^ with an onset within 0-10 days before the date of sample collection. We chose the matching factors that were predictors of vaccination and SARS-CoV-2 infection (Supplementary Figures 3-6), and excluded individuals with missing matching factors. Due to the limited study size, we chose a small set of matching factors to balance the ability to reduce bias and to enrol sufficient case-control pairs to achieve desired power. We chose to categorize age as <30, 30-60, and ≥60 years because the proportion of RT-PCR tests that were positive appeared to be consistent within these age bands, reflecting a similar risk for infection in this group, albeit possibly differential healthcare utilization (Supplementary Figure 3). Upon identification of each case, a control was selected at random from a list of eligible matching controls, such that each eligible control was matched to at most one case.

### Statistical analysis

For the early analysis, we defined vaccination as having received at least one dose of the two-dose CoronaVac schedule, with the first dose administered ≥14 days before the sample collection date for their RT-PCR test. We pre-specified the early analysis of the effectiveness of at least one dose in the study protocol and used the O’Brien Fleming alpha-spending method to calculate an adjusted critical p-value of 0.0492.^26^ For the planned analysis of effectiveness after two doses, we defined vaccination in three categories: having received a single dose administered ≥14 days before the sample collection date for their RT-PCR test; having received two doses, with the second dose administered 0-13 days before the sample collection date for their RT-PCR test; and having received two doses, with the second dose administered ≥14 days before the sample collection date for their RT-PCR test. The reference group for vaccination status was individuals who had not received a first vaccine dose by the date of sample collection.

Finally, we evaluated the exposure of receiving the first vaccine dose from 0 to 13 days before the sample collection date, a period where the vaccine likely has no or limited effectiveness.^14,15^ A non-null association between this exposure and SARS-CoV-2 infection risk compared to those who had not received a first vaccine dose by the date of sample collection may serve as an indicator of unmeasured confounding.

Analyses of the primary outcome of symptomatic SARS-CoV-2 infection included case-control pairs who had symptomatic illness 0-10 days before the time of testing. Analyses of the secondary outcome of any SARS-CoV-2 infection included additional case-control pairs who did not have symptomatic illness before or at the time of testing.

We used conditional logistic regression to estimate the odds ratio (OR) of vaccination among cases and controls.^27^ 1-OR provided an estimate of vaccine effectiveness under the assumptions of a test-negative design.^28^ We included as covariates in the adjusted model: sex, occupation category, self-reported race/skin colour, number of previous healthcare interactions from the beginning of the pandemic (March 2020) to the start of the study, and a SARS-CoV-2 infection, defined as a positive RT-PCR or antigen detection test, from the beginning of the pandemic (March 2020) to the start of the study period. A missing indicator was incorporated to address missing information on occupation category or race. We adjusted for the possible effect of COVID-19-associated comorbidities^29^ in a separate sensitivity analysis due differential completeness for this covariate between cases and controls. Finally, we performed a sensitivity analysis excluding any case-control pairs including at least one individual with a previous SARS-CoV-2 infection.

### Power calculation and timing of analyses

After generating matched case-control pairs for each pre-specified analysis and before performing the analysis, we simulated the power of the data set to identify a given vaccine effectiveness (see the Supplementary File, p5 for details). After extracting the surveillance databases on 1 April, 2021, we determined that the power of the early analysis to identify a vaccine effectiveness of 60% of at least one dose was 92.7%. On 20 April, 2021, we determined that the power of the two-dose analysis to identify a vaccine effectiveness of 70% comparing those with two doses ≥14 days after the second dose to those who had not received a vaccine was 80.0%.

All analyses were done in R, version 4.0.2.

## Results

### Second COVID-19 epidemic and vaccination campaign among HCWs in Manaus

Among 68,808 HCWs that were employed in the city’s healthcare facilities, 67,718 could be linked to surveillance databases (Figure 2). In this population, 3,445 cases of SARS-CoV-2 infection were reported from 1 October 2020 to 13 April 2021 (Figure 1). Among the 3,445 cases, 2,559 and 886 were associated with and without, respectively, COVID-19 symptoms. The municipal vaccination campaign was initiated on 19 January, 2021 and as of 13 April, 2021, has administered first and second vaccine doses to 55,584 (82%) and 50,029 (74%), respectively, of the 67,718 HCWs.

### Study population for the early at-least-one-dose analysis

Among the 67,718 HCWs, 53,176 were eligible for inclusion in the early analysis (Figure 2). Of the 53,176 HCWs, 1,752 and 904 received RT-PCR testing during the study period of 19 January to 25 March, 2021 who respectively, did or did not report a symptomatic illness at the time of testing. Among the 1,823 and 974 tests performed for HCWs with and without symptomatic illness, respectively, 564 (31% of 1,823) and 212 (22% of 974), respectively, were positive. Through matching, we selected 780 HCWs with 786 RT-PCR tests to establish 393 case-control pairs with symptomatic illness and 266 HCWs with 270 RT-PCR tests to establish 135 pairs without symptomatic illness.

### Study population for the two-dose analysis

Among the 67,718 HCWs, 53,153 were eligible for inclusion in the two-dose analysis (Figure 2). Of the 53,153 HCWs, 1,907 and 1,038 individuals received RT-PCR testing during the study period of 19 January to 13 April, 2021 who respectively, did or did not report a symptomatic illness at the time of testing. Among the 1,985 and 1,116 tests performed for HCWs with and without symptomatic illness, respectively, 590 (30%) and 218 (20%), respectively, were positive. Through matching, we selected 829 HCWs with 836 RT-PCR tests to establish 418 case-control pairs with symptomatic illness and 272 HCWs with 276 RT-PCR tests to establish 138 pairs without symptomatic illness. The characteristics of HCWs matched, unmatched, and not eligible for case or control selection, are shown in Supplementary Table 1, the timing of pair enrolment during the study period is shown in Supplementary Figure 7 (early analysis) and Supplementary Figure 8 (two-dose analysis), and the distribution of discordant pairs is shown in Supplementary Table 2 (early analysis) and Supplementary Table 3 (two-dose analysis).

Table 1 shows the distribution of characteristics between symptomatic cases and controls selected for the early analysis (left) and two-dose analysis (right). The proportion of females was lower among the cases. The proportion who had a positive SARS-CoV-2 RT-PCR or antigen test prior to the study period was small, but higher among controls than cases (3.1 vs 6.9%). These differences suggest potential for confounding in the unadjusted analysis. Of the 556 cases in the two-dose analysis, 10.6% (59) required hospitalisation for their SARS-CoV-2 infection. Among those who had received at least one dose of vaccine before the RT-PCR sample collection date, the average time from first dose to sample collection date was 13 days (IQR 7-26) among cases and 17 days (IQR 8-28) among controls (Supplementary Figure 9). Among those who had received both vaccine doses before the RT-PCR sample collection date, the average time from second dose to sample collection date was 14 days (IQR 7-27) among cases and 14 days (IQR 7-21) among controls (Supplementary Figure 9). Supplementary Table 4 shows the distribution of characteristics between cases and controls without symptomatic illness at the time of testing.

**Table 1:** Comparison of symptomatic cases and controls

|  | Early at-least-one-dose analysis |  | Two-dose analysis |  |
| --- | --- | --- | --- | --- |
| Characteristics | Cases (n=393) | Controls (n=393) | Cases (n=418) | Controls (n=418) |
| Vaccination |  |  |  |  |
| Not vaccinated | 231 (59%) | 238 (61%) | 238 (59%) | 255 (61%) |
| One dose (first dose 0-13 days previously) | 89 (23%) | 59 (15%) | 95 (23%) | 58 (19%) |
| One dose (first dose $\geq 14$ days previously) | 73 (19%) | 96 (24%) | 41 (10%) | 52 (12%) |
| Two doses (second dose 0-13 days previously) |  |  | 21 (5%) | 26 (6%) |
| Two doses (second dose $\geq 14$ days previously) | | | 23 (6%) | 27 (7%) |
| Age (years, mean (SD)) | 43.3 (9.5) | 42.7 (9.4) | 43.2 (9.5) | 42.6 (9.4) |
| Female sex | 276 (70%) | 313 (80%) | 291 (70%) | 332 (79%) |
| Self-reported race/skin colour* |  |  |  |  |
| Amarela/Yellow | 52 (13%) | 70 (18%) | 59 (14%) | 76 (18%) |
| Preta/Black | 5 (1%) | 3 (1%) | 5 (1%) | 3 (1%) |
| Pardo/Brown | 237 (60%) | 229 (58%) | 252 (60%) | 242 (58%) |
| Branca/White | 81 (21%) | 73 (19%) | 82 (20%) | 77 (18%) |
| Missing | 18 (5%) | 18 (5%) | 20 (5%) | 20 (5%) |
| Occupation category |  |  |  |  |
| Administrative | 108 (28%) | 90 (23%) | 115 (28%) | 92 (22%) |
| Clinician | 17 (4%) | 8 (2%) | 18 (4%) | 9 (2%) |
| Nurse/nurse technician | 152 (39%) | 170 (43%) | 161 (39%) | 181 (43%) |
| Other health professional | 41 (10%) | 44 (11%) | 43 (10%) | 47 (11%) |
| Other health associated | 68 (17%) | 68 (17%) | 72 (17%) | 73 (18%) |
| Missing | 7 (2%) | 13 (3%) | 9 (2%) | 16 (4%) |
| Number of healthcare encounters<sup>†</sup> | 0.81 (1.09) | 0.83 (1.14) | 0.81 (1.08) | 0.79 (1.06) |
| Prior positive SARS-CoV-2 test<sup>†‡</sup> | 11 (3%) | 26 (7%) | 13 (3%) | 29 (7%) |

\*Race/skin colour as defined by the Brazilian national census bureau (Instituto Nacional de Geografia e Estatísticas) https://biblioteca.ibge.gov.br/visualizacao/livros/liv63405.pdf
<sup>†</sup>From March, 2020 to the start of the study on 19 January, 2021.
<sup>‡</sup> Defined as SARS-CoV-2 RT-PCR or antigen detection.

### Early at-least-one-dose analysis

After adjusting, vaccination with at least one CoronaVac dose was associated with a 0.51-fold reduction (adjusted VE, 49.4%, 95% CI 13.2 to 71.9) in the odds of symptomatic SARS-CoV-2 infection during the period 14 days or more after receiving the first dose (Table 2). Of note, the odds of symptomatic SARS-CoV-2 infection was increased (aOR 1.66, 95% CI 1.07 to 2.57) amongst vaccinated HCWs in the period 0-13 days after receiving the first vaccine dose when compared with HCWs who did not receive the vaccine. Female sex (aOR 0.56, 95% CI 0.38 to 0.80) and a positive SARS-CoV-2 RT-PCR or antigen test in the pre-study period (aOR 0.38, 95% CI 0.17 to 0.87) were associated with a reduced odds of symptomatic SARS-CoV-2 infection. Estimated vaccine effectiveness of at least one dose against all SARS-CoV-2 infection during the period 14 days or more after receiving the first dose was 35.1% (95% CI -6.6 to 60.5) (Supplementary Table 5, aOR 0.65, 95% CI 0.40 to 1.07).

**Table 2:** Vaccine effectiveness against symptomatic SARS-CoV-2 infection, from the early analysis, with at least one dose and at least 14 days after administration of the first dose, and the two-dose analysis, with effectiveness by dose

|  | Early at-least-one-dose analysis |  | Two-dose analysis |  |
| --- | --- | --- | --- | --- |
|  | OR<br>(95% CI) | p-value | OR<br>(95% CI) | p-value |
| Unadjusted Analysis |  |  |  |  |
| 1 dose: 0-13 days after 1<sup>st</sup> vaccine dose vs. unvaccinated* | 1.61 (1.07-2.44) | 0.02 | 1.89 (1.25-2.84) | <0.001 |
| 1 dose: ≥14 days after 1<sup>st</sup> vaccine dose vs. unvaccinated* | 0.56 (0.32-0.95) | 0.03 | 0.68 (0.38-1.21) | 0.19 |
| 2 doses: 0-13 days after 2<sup>nd</sup> vaccine dose vs. unvaccinated* |  |  | 0.62 (0.30-1.31) | 0.22 |
| 2 doses: ≥14 days after 2<sup>nd</sup> vaccine dose vs. unvaccinated* |  |  | 0.60 (0.25-1.40) | 0.23 |
| Adjusted analysis |  |  |  |  |
| 0-13 days after 1<sup>st</sup> vaccine dose vs. unvaccinated* | 1.66 (1.07-2.57) | 0.02 | 2.07 (1.34-3.21) | <0.001 |
| ≥14 days after 1<sup>st</sup> vaccine dose vs. unvaccinated* | 0.49 (0.28-0.87) | 0.01 | 0.62 (0.34-1.14) | 0.13 |
| 2 doses: 0-13 days after 2<sup>nd</sup> vaccine dose vs. unvaccinated* |  |  | 0.46 (0.20-1.04) | 0.06 |
| 2 doses: ≥14 days after 2<sup>nd</sup> vaccine dose vs. unvaccinated* |  |  | 0.63 (0.26-1.53) | 0.31 |
| Female sex | 0.56 (0.38-0.8) | <0.001 | 0.54 (0.37-0.78) | <0.001 |
| Self-reported race/skin colour<sup>†</sup> |  |  |  |  |
| Amarela vs. Pardo | 0.77 (0.50-1.20) | 0.25 | 0.81 (0.53-1.24) | 0.33 |
| Preta vs. Pardo | 1.89 (0.41-8.80) | 0.41 | 2.06 (0.44-9.56) | 0.36 |
| Branca vs. Pardo | 1.18 (0.80-1.75) | 0.41 | 1.09 (0.74-1.61) | 0.66 |
| Occupation category |  |  |  |  |
| Administrative vs. Nurse | 1.10 (0.74-1.62) | 0.65 | 1.16 (0.79-1.71) | 0.45 |
| Clinical vs. Nurse | 2.16 (0.82-5.69) | 0.12 | 1.95 (0.76-4.93) | 0.16 |
| Other health associated vs. Nurse | 0.95 (0.60-1.52) | 0.84 | 0.95 (0.60-1.50) | 0.83 |
| Other health professional vs. Nurse | 0.89 (0.53-1.51) | 0.67 | 0.92 (0.55-1.56) | 0.76 |
| Prior healthcare encounters<sup>‡</sup> |  |  |  |  |
| 1-3 vs. 0 | 1.03 (0.75-1.41) | 0.87 | 1.09 (0.79-1.49) | 0.60 |
| ≥4 vs. 0 | 1.50 (0.59-3.87) | 0.40 | 2.92 (1.00-8.52) | 0.05 |
| Prior positive SARS-CoV-2 test<sup>‡**</sup> | 0.38 (0.17-0.87) | 0.02 | 0.36 (0.17-0.79) | 0.01 |

\*At date of index sample collection for cases and controls
<sup>†</sup>Race/skin colour as defined by the Brazilian national census bureau (Instituto Nacional de Geografia e Estatísticas) https://biblioteca.ibge.gov.br/visualizacao/livros/liv63405.pdf
<sup>‡</sup>From March, 2020 to the start of the study on 19 January, 2021
\*\* Defined as SARS-CoV-2 RT-PCR or antigen detection

### Two-dose analysis

Upon accruing more case-control pairs, we performed an analysis of vaccine effectiveness by dose. After adjusting, vaccination with two doses of CoronaVac was not associated (adjusted VE 37.1%, 95% CI -53.3 to 74.2) with a reduction in the odds of symptomatic SARS-CoV-2 infection during the period 14 days or more after receiving the second dose. As in the early analysis, vaccination 0-13 days before sample collection was associated with increased odds of symptomatic SARS-CoV-2 infection, while female sex and prior positive SARS-CoV-2 viral test were associated with reduced odds of symptomatic SARS-CoV-2 infection (Table 2). Estimated vaccine effectiveness of two doses against all SARS-CoV-2 infection during the period 14 days or more after receiving the second dose was 37.9% (95% CI -46.4 to 73.6) (Supplementary Table 5, aOR 0.62, 95% CI 0.26 to 1.46). Estimated vaccine effectiveness of two doses against asymptomatic SARS-CoV-2 infection during the period 14 days or more after receiving the second dose was 100% (95% CI undefined) (Supplementary Table 6), but this analysis was very underpowered.

For primary and secondary outcomes, estimates for vaccine effectiveness and significant covariates were similar when adjusting for presence of one or more underlying comorbidities (Supplementary Table 7). In a sensitivity analysis excluding pairs where either individual had a previous positive SARS-CoV-2 test, we estimated effectiveness of two doses to be 56.2% (95% CI -23.0 to 84.4).

## Discussion

Here, we provide suggestive but inconclusive evidence for the effectiveness of CoronaVac in the setting of widespread Gamma variant transmission. An RCT of CoronaVac in Brazil^16^ reported efficacy of 50.7% (95% CI 35.6 to 62.2) against symptomatic COVID-19 (score ≥2 on the WHO Clinical Progression Scale),^30^ and an RCT from Turkey provided consistent evidence for the efficacy of this vaccine.^31^ However, these trials were conducted prior to the emergence of the Gamma variant.^31^ Several preliminary studies have assessed the effectiveness of CoronaVac in other populations, including elderly individuals in São Paulo State,^32^ HCWs in a single centre in São Paulo,^33^ elderly age groups in Brazil,^34^ and the general population in Chile.^31^ More evidence is needed from observational studies conducted in Brazil and other countries where the Gamma variant or other lineages are circulating.

In an effort to translate results of public health importance as early as possible, we performed a planned analysis of vaccine effectiveness following at least one dose and reported estimated effectiveness of 49.4% (95% CI 13.2 to 71.9) against symptomatic COVID-19, starting 14 days after administration of the first dose, in healthcare workers from Manaus. However, a planned subsequent analysis of effectiveness by dose failed to show strong effectiveness of two doses, with estimated effectiveness of 37.1% (95% CI -53.3 to 74.2). Although the early results are promising, the evidence from this study is mixed. Studies will be required which have larger sample sizes and evaluate other populations, particularly those with lower seroprevalence, to assess the real-world effectiveness of CoronaVac and support the efficacy shown in RCTs. In addition, we did not achieve sufficient power for pre-specified analyses that were proposed to evaluate vaccine effectiveness against severe outcomes and against COVID-19 in elderly individuals in Manaus. Finally, the low estimated effectiveness against all infection suggests that indirect effects may be low in this setting, and that high vaccine coverage must be achieved to maximize population-level impacts of vaccination.

Our analysis of effectiveness by dose was limited in several ways. SARS-CoV-2 seroprevalence in Manaus was likely high prior to the vaccination campaign.^23,24^ Prior natural infection may have conferred protection to unvaccinated individuals, which in turn may lead to underestimation of the VE estimate among seropositive individuals. If vaccine uptake were lower among those previously infected, this would exacerbate such a downward bias. The proportion with a previous positive RT-PCR or antigen test was in fact slightly higher among vaccinated individuals (5.3% vs. 3.6%), possibly representing higher access to healthcare. Due to the limited access to RT-PCR and antigen detection testing in the city, this variable does not fully control for this source of confounding, nor did we have sufficient power to explore the effect modification of vaccine effectiveness by previous infection. A sensitivity analysis suggested higher vaccine effectiveness among individuals without evidence of a previous infection, although the precision remained low. For studies of COVID-19 vaccines not containing the N protein, testing for antibodies against the N protein to determine infection history could be an important part of study design to increase validity.

The precision of the analysis of effectiveness of the two-dose schedule was low. We had powered this analysis to detect effectiveness of 70% after two doses, but it may be that true effectiveness is lower in this population. Several studies found that vaccination in individuals with a previous infection elicits a strong and rapid immune response,^35,36^ as well as cross-neutralizing antibody response to the Gamma variant.^7^ Overall immunogenicity following a single dose in this population may thus be higher than in a low-seroprevalence population. Conversely, the effectiveness of the second dose relative to the first dose may be similar. As median time from second dose to sample collection date was 14 days among those who received two doses, we were unable to assess any changes in effectiveness over longer follow-up times, and our estimate may represent effectiveness at the peak of antibody titre following vaccination.

We could not directly address whether CoronaVac was effective against the Gamma variant as SARS-CoV-2 samples from HCW cases were not routinely genotyped. However, the study was conducted at the epicentre for Gamma variant emergence and during an epidemic when surveillance of the general population identified the variant in 66% of genotyped samples. It seems plausible that vaccination with CoronaVac conferred a level of protection against the Gamma variant and that our estimates reflect VE in the real-world setting of high Gamma variant transmission, but such a finding should be strengthened by further studies and examined for other vaccine platforms.

We addressed multiple potential sources of bias in this observational setting. The use of a test-negative design allowed for control of healthcare-seeking behaviour among study participants, albeit with limits. For the analysis of the primary outcome, we restricted cases and controls to patients with evidence of any symptoms proximal to the time of testing. This restriction reduces the risk of outcome misclassification, as sensitivity of PCR tests is high in this time period. However, exposure to SARS-CoV-2 likely differs between those presenting with different symptoms, in particular for those without symptoms. We were unable to perform additional matching with detailed symptom data and cannot exclude the possibility of differential test-seeking or exposure between vaccinated and unvaccinated individuals with varying symptomatology,^37^ nor of misclassification due to false negative PCR tests. However, a strength of the surveillance system in Manaus was the large source of individual-level data that allowed us to match on a number of variables in order to reduce confounding.

Our estimates may be subject to unmeasured and residual confounding, as unvaccinated individuals receiving PCR tests may have different risk of SARS-CoV-2 infection than vaccinated individuals for reasons unrelated to vaccination. We addressed this possibility by evaluating the risk associated with being vaccinated 0-13 days before testing, when vaccination is likely to be ineffective or have reduced effectiveness.^14,15^ There are several possible explanations for the observed positive association: HCWs prioritised for vaccination could have been at higher risk of SARS-CoV-2 exposure than those who were vaccinated later; HCWs were unlikely to receive a positive test in the time immediately preceding their vaccination (reverse causation); or recently vaccinated individuals only sought testing for more severe symptoms, which were more likely to be due to SARS-CoV-2. This bias indicator is likely due to a combination of time-invariant and time-varying differences in exposure risk or testing behaviour between vaccinated and unvaccinated HCWs, as suggested in other studies.^38,39^ It suggests that confounding contributed to underestimation of the VE in the analysis of at-least-one-dose and two-dose schedules.

We observed a strong protective effect of previous positive RT-PCR or antigen detection test (aOR 0.36, 95% CI 0.17 to 0.79) and of female sex (aOR 0.53, 95% CI 0.37 to 0.77), consistent with other studies.^40,41^ The association between previous positive SARS-CoV-2 test and infection is likely an underestimate of the protection from infection, as many without a positive test were truly infected.

Evidence from this study is mixed and highlights that as vaccination campaigns with CoronaVac continue in Brazil and other countries, non-pharmaceutical interventions are necessary to reduce transmission, morbidity, and mortality in the population.

## Supporting information

STROBE Checklist

## Data Availability

Deidentified databases as well as the R codes will be deposited in the repository https://github.com/juliocroda/VebraCOVID-19

## Contributors

MDTH and OTR share co-first authorship. MSST and SBO were responsible for linkage, cleaning and de-identifying the databases. MDTH, OTR, DATC, JRA, AIK and JC participated in the design and concept of the study and designed the data analysis. MDTH and OTR wrote the first version of the manuscript. WNA, MA, RS, AMS, BCA, SHHF, CFC supervised the study. All authors participated in data interpretation, revised the manuscript, and approved the final version of the manuscript. MSST and SBO have verified the underlying data.

## Data sharing

De-identified databases as well as the R codes will be deposited in the repository https://github.com/juliocroda/VebraCOVID-19

## Acknowledgements

We are grateful for Pan American Health Organization’s support to the Fundação de Vigilância em Saúde from Amazonas State and the Municipal Health Secretary of Manaus in making the databases available for analysis. JC and AS are supported by the Oswaldo Cruz Foundation (Edital Covid-19 – resposta rápida: 48111668950485). OTR is funded by a Sara Borrell fellowship (CD19/00110) from the Instituto de Salud Carlos III. OTR acknowledges support from the Spanish Ministry of Science and Innovation through the Centro de Excelencia Severo Ochoa 2019-2023 Program and from the Generalitat de Catalunya through the CERCA Program. We thank Julio Horta for automating database linkage.

