## Supplementary Appendix for "Effectiveness of CoronaVac among healthcare workers in the setting of high SARS-CoV-2 Gamma variant transmission in Manaus, Brazil: A test-negative case-control study"

### Effectiveness of an inactivated SARS-Cov-2 vaccine amongst healthcare workers in the setting of high P.1 variant transmission in Brazil

#### SUPPLEMENTARY MATERIALS

**Table of Contents:**


- pp 1-7: Study protocol
- p 8: Supplementary Figure 1
- p 9: Supplementary Figure 2
- p 10: Supplementary Figure 3
- p 11: Supplementary Figure 4
- p 12: Supplementary Figure 5
- p 13: Supplementary Figure 6
- p 14: Supplementary Figure 7
- p 15: Supplementary Figure 8
- p 16: Supplementary Figure 9
- p 17: Supplementary Table 1
- p 18: Supplementary Table 2
- p 18: Supplementary Table 3
- p 19: Supplementary Table 4
- p 20: Supplementary Table 5
- p 22: Supplementary Table 6


**PROTOCOL**

##### Evaluation of Vaccine Effectiveness in Brazil against COVID-19 (VEBRA-COVID): A Test-Negative Case-Control Study Protocol

Version: 01.1 / April 3rd 2021

#### I. Background

Since the emergence of severe acute respiratory virus coronavirus 2 (SARS-CoV-2), Brazil has experienced one of the world's highest incidence and mortality rates in the world, with over 10 million reported infections as of the end of February 2021.[1–3] Of grave concern is the resurgence of COVID-19 cases and hospitalizations observed in the city of Manaus, despite high estimated seroprevalence.[4,5] One hypothesis for this resurgence is loss of immunity to the P.1 Variant of Concern (VOC), which was first detected in Manaus on Jan 12, 2021, and now consists the majority of new infections, including all genomes sequenced in this city in February 2021.[6–8] This lineage has accrued mutations associated with decreased neutralization,[9,10] and has since spread throughout Brazil.

The rapid development of novel vaccines against COVID-19 allowed countries to start vaccine distribution programs within a year of the identification of the novel virus. Among the first vaccines to be developed was Sinovac's CoronaVac vaccine.[11,12] Phase III trials were conducted in Turkey and Brazil, and results were announced on February 5, 2021, in which effectiveness after 14 days following vaccination with 2 doses of vaccine was reported to be 50.65% for all symptomatic cases of COVID-19, 84% for cases requiring medical attention, and 100% for hospitalized, severe, and fatal cases.[13] CoronaVac was approved for emergency use on 17 January in Brazil, and used to vaccinate healthcare workers and the general population, beginning with the oldest age groups, on 19 January 2021. AstraZeneca-Oxford's ChAdOx1 vaccine was approved on the same day and was administered beginning on 23 January 2021.

As vaccine programs continue, there has been much interest in estimation of vaccine effectiveness through observational studies, and specifically in settings where VOC are circulating. Such studies have advantages over clinical trials, including increased size and follow-up time, and reduced cost. However, as vaccinated and unvaccinated individuals are likely different in their SARS-CoV-2 risk and healthcare access, these studies must address bias through design and analysis. Several studies have demonstrated the effectiveness of COVID-19 vaccines against infection caused by the B.1.1.7 variant.[14] However, large-scale real-world investigations on vaccine effectiveness have not been conducted in regions where the P.1 variant is prevalent.

We propose a test-negative case-control study[15,16] of healthcare workers (HCWs) from the city of Manaus to evaluate the effectiveness of vaccines in preventing COVID-19 in a setting of widespread P.1 VOC transmission.[7] Manaus was selected as the site for the HCW study since it was the first city to aggressively vaccinate HCWs in response to the P.1 epidemic. The study will be limited to evaluating the effectiveness of CoronaVac since 97% of vaccinated HCWs in Manaus received this vaccine. We will expand the study population as additional age groups become eligible for vaccination. Furthermore, we expect that additional vaccines will be approved and will evaluate their effectiveness. We will therefore continue to amend the protocol and its objectives accordingly to address these new questions.

#### II. Objectives

1. To estimate the effectiveness of CoronaVac against symptomatic SARS-CoV-2 infection amongst healthcare workers from the city of Manaus.

#### III. Methods

**1. Study Design**: We will conduct a retrospective matched case-control study, enrolling cases who test positive for SARS-CoV-2 and controls who test negative for SARS-CoV-2 amongst HCWs (Section 3) and the general population (Section 4) as of the day that the COVID-19 vaccination campaign was initiated at the study sites. The study will evaluate vaccine effectiveness on the primary outcome of symptomatic SARS-CoV-2 infection and secondary outcome of SARS-CoV-2 RT-PCR test positivity regardless of symptoms. We will identify cases and matched controls by extracting information from health surveillance records and ascertain the type and data of vaccination by reviewing the state COVID-19 vaccination registry. In this design, the odds ratio of vaccination comparing cases and controls estimates the direct effect of vaccination on the disease outcome. We will perform interim analyses aimed at evaluating the effectiveness of receiving at least one vaccine dose and a final analysis that will evaluate the effectiveness of completing the approved vaccine series. In a separate analysis, we will assess the association between vaccination and hospitalization and/or death among individuals who have tested positive for SARS-CoV-2.

**2. IRB and Ethics Statement**: The protocol has been submitted to the Ethical Committee for Research of Federal University of Mato Grosso do Sul (CAAE: 43289221.5.0000.0021). The work of investigators at the University of Florida, Yale University, Stanford University, and Barcelona Institute for Global Health was conducted to inform the public health response and was therefore covered under Public Health Response Authorization under the US Common Rule.

**3. Study Details**

Study Site: Manaus (3°5′S, 60°W) is the capital of the state of Amazonas, the major urban metropolis in the middle of the Amazon jungle, and a major river port for seafaring vessels. In 2020, Manaus, with an estimated population of 2,219,580 inhabitants, reported 144,767 COVID-19 cases (cumulative incidence: 6,522 per 100,000 population) and 7,605 deaths (cumulative mortality: 342 per 100,000 population). Manaus has 40 Family Health (*Plano Saúde da Familia*) teams, 53 primary health care centers and 15 other health units under the responsibility of the Secretariat of Health of Manaus, and 20 private or public hospitals. The Municipal Secretary of Health of Amazonas initiated its COVID-19 vaccination campaign on 19 January 2021 and is administering two vaccines, CoronaVac and ChAdOx1. CoronaVac has been used in >97% of the vaccinations of HCWs.

Data Sources and Integration: The overall approach will be to: 1) Identify the cohort of all HCW from Manaus from *state HCW registries*; 2) Identify eligible cases and controls from the cohort who test positive and negative, respectively, from the *state laboratory testing registry* of public health laboratory network; 3) Determine vaccination status from *municipal vaccination registries*; and 4) Extract information from *national healthcare and surveillance databases* that will be used to define outcomes, match controls to cases, determine vaccination status, serve as covariates for post-stratification and provide a source for cross-validation of information from databases. Data sources will include:

- SES-AM HCW registry
- National health plan registry of users (**CADSUS**)
- National laboratory testing registry (**GAL**) of the network of public health laboratories
- Municipal COVID-19 vaccination registry
- National surveillance database of severe acute respiratory illnesses (**SIVEP-Gripe**) created by Ministry of Health Brazil in 2009
- National surveillance system of suspected cases of COVID-19 (**e-SUS**) from mild to moderate "influenza like illness", created by the Ministry of Health Brazil in 2020
    - e-SUS includes information from healthcare telemonitoring, whereby teams of healthcare practioners make daily telephone calls, assess symptoms, identify signs of severity and triage patients to healthcare facilities.
- National mortality registry (**SIM**) from the Ministry of Health

We will build a parent database using the mySQL language and integrated individual datasets using an application programming interface (API), which was developed using ElasticSearch. Table 1 in the Supplementary material lists the variables extracted from datasets and incorporated in the parent database. The database will be updated on a weekly basis.

We will use CPF numbers (Brazilian citizens' unique identifier code) to integrate datasets. For those entries missing CPF in SEMSA, we will perform a probabilistic record linkage between SEMSA and CADSUS (registration database of users of the public universal health system [SUS] in Brazil). For the probabilistic method we will use the Reclink III software,[17] and consider sex, the phonetic code of the first and last name and the phonetic code of the first name of the mother as blocking variables. We will compare the similarity of the name, mother's name with a threshold of >85% similarity, and date of birth a threshold of >65% similarity. All pairs identified by the probabilistic method will be manually reviewed and revised.

Some variables were reported in multiple data sources. To define a single variable, we drew from each database with priority given to databases that were more complete, reliable, and up-to-date. We will choose age from the data sources in the following order: CADSUS, SEMSA, GAL, e-SUS, SIVEP-Gripe. We will define neighborhoods (*bairro*) by extracting information on CEP (Brazilian zipcode) and transforming them to neighborhoods or directly extracting information on neighborhoods from data sources in the following order: CADSUS (CEP), CADSUS (*bairro*), SEMSA (CEP), and e-SUS (CEP).

Study Population

*Inclusion criteria*:

- Healthcare worker as defined by the SEMSA registry,
- Has a residential address within the city of Manaus,
- Age ≥ 18 years before 19 January 2021,
- With complete information, which is consistent between databases, on age, sex, and residential address defined by CEP (zip code)
- With complete and consistent vaccination status and dates.

*Exclusion criteria*:

- Not a healthcare worker as defined by the SEMSA/SES-AM registry,
- Does not have a residential address within the city of Manaus,
- Aged < 18 years before 19 January 2021,
- With missing or inconsistent information on age, sex, or residential address defined by CEP (zip code)
- With incomplete or inconsistent vaccination status or dates.

Case definition and eligibility: We will use information from integrated GAL/SIVEP-Gripe/e-SUS databases to identify eligible cases. Cases are defined as eligible members of the study population (as defined above, Study Population) who:

- Had a sample with a positive SARS-CoV-2 RT-PCR, which was collected between January 19, 2021 and 7 days prior to database extraction of information
- Did not have a positive RT-PCR test in the preceding 90 day period,
- Have complete and consistent data on SARS-CoV-2 PCR test result,
- Did not receive a dose of ChAdOx1 vaccine before the date of respiratory sample collection.

Control definition and eligibility: We will use integrated GAL/SIVEP-Gripe/e-SUS databases to identify eligible controls. Controls are defined as eligible members of the study population who:

- Had a sample with a negative SARS-CoV-2 RT-PCR result, which was collected after January 19, 2021,
- Did not have a subsequent positive PCR test in the following 7-day period
- Have complete and consistent data on SARS-CoV-2 PCR test result,
- Did not receive a dose of ChAdOx1 vaccine before the date of respiratory sample collection.

Matching: Test-negative controls will be matched 1:1 to the cases. Matching factors will include variables that are anticipated to be causes of the likelihood of receiving the vaccine, risk of infection and likelihood of receiving PCR testing for SARS-CoV-2 (i.e. healthcare access and utilization) (see Figures 1-3):

- Symptomatic illness status at time of testing, defined as the presence or absence of one or more reported COVID-19 related symptom with onset within 0-10 days before the date of their positive and negative PT-PCR test, respectively, for case and controls,
- Residential address (neighborhood [*bairro*], which is identified based on the first 5 digits of 8 digit CEP),
- Age (categorized as <30, ≥30 and <60, and ≥60 years; Figure 3 shows similar rates of testing positive for individuals aged 30-60),
- Window of ±3 days between collection of RT-PCR positive respiratory sample for cases and collection of RT-PCR negative respiratory sample for controls. If the date of respiratory sample collection is missing, the date of notification of testing result will be used.

We chose the matching factors to balance the ability to reduce bias and to enroll sufficient case-control pairs. We chose to categorize age as <30,30-60, and ≥60 years because the proportion of RT-PCR tests that were positive appeared to be fairly constant in the middle age band, reflecting a similar risk for infection in this group, albeit possibly differential healthcare utilization. We observed a negative correlation between the proportion of positive RT-PCR tests in a neighborhood and the average time until receipt of first dose, possibly implying a positive correlation between access to testing and access to vaccination. Finally, as incidence decreased over the study period as vaccine coverage increased, time was a clear confounder, and matching was considered an efficient way to address this bias.

We will use the standard algorithms to conduct matching which include: 1) setting a seed, 2) locking the database, 4) creating a unique identifier for matching after random ordering, 5) implementing exact matching based on matching variables, sampling controls at random if more than one available per case within strata.

An individual who fulfils the control definition and eligibility and later has a sample tested that fulfils the case definition and eligibility can be included in the study as both a case and a control. An individual who fulfils the control definition for multiple different sample collection dates can be included in the study as a control for each collection date, up to a maximum of three times.

Exposure definition: CoronaVac vaccination in the following stratifications:

- Received the first vaccine dose, and not having received a second dose, in the following time periods relative to sample collection for their PCR test:
    - 0-13 days
    - ≥14 days
- Received the second dose in the following time periods relative to sample collection for their PCR test:
    - 0-13 days
    - ≥14 days

Statistical Analyses: We will evaluate the effectiveness of CoronaVac for the following SARS-CoV-2 infection outcomes:

- Primary: Symptomatic COVID-19, defined as one or more reported COVID-19 related symptom with onset within 0-10 days before the date of their positive RT-PCR test
- Secondary:
    - SARS-CoV-2 RT-PCR test positivity
    - COVID-19 associated hospitalization within 14 days of the symptom onset
    - COVID-19 associated death within 28 days of symptom onset

We will evaluate vaccine effectiveness for the primary outcome and the secondary outcome of test positivity in case control analyses according to the test-negative design. Table 2 shows a list of all planned analyses in the test-negative design. The test-negative design may introduce bias when evaluating outcomes of hospitalizations and deaths during an epidemic. We will therefore perform survival analyses of HCWs who test positive to evaluate the association of vaccination status and the risk for hospitalization and death after infection.

***Case-control analysis***: Analyses of the primary outcome will be restricted to case and control pairs who are matched based on the presence of a COVID-19 related symptom before or at the time of testing. Analyses of the secondary outcomes of test positivity will include additional case and control pairs who are matched based on the absence of a COVID-19 related symptom before or at the time of testing.

We will use conditional logistic regression to estimate the odds ratio (OR) of vaccination among cases and controls, accounting for the matched design, where 1-OR provides an estimate of vaccine effectiveness under the standard assumptions of a test-negative design. The reference group will be individuals who have not received a first dose of CoronaVac by the date of respiratory sample collection. Date of notification of the testing result will be used if the date of respiratory sample collection is missing. To evaluate potential biases and the timing of vaccine effectiveness after administration, we will evaluate the windows of vaccination status corresponding to 0-13 days and ≥14 days after the first dose and 0-13 days and ≥14 days after the 2nd dose.

We will include the following covariates in the adjusted model, which we hypothesize are predictive of vaccination, the risk of SARS-CoV-2 infection and COVID-19 severity and healthcare access and utilization:

- Age as continuous variable
- Sex
- Occupation category
- Self-reported race/skin color
- Number of previous entries in e-SUS or SIVEP-Gripe surveillance databases
- Evidence of prior SARS-CoV-2 infection (defined as positive PCR test, antigen test or rapid antibody test)

Although data on comorbidities is available through e-SUS and SIVEP-Gripe, this data may have different degrees of missingness between databases and between cases and control groups. Adjusting for comorbidities using complete case data will likely introduce bias. We will explore the feasibility of multiple imputation of comorbidity in a sensitivity analysis. Additional sensitivity analyses will evaluate potential effect modification of the vaccine effectiveness by history of a positive RT-PCR, antigen or serological test result prior to the vaccination campaign.

***Survival analysis of h*****ospitalization and death**: We will perform proportional survival analyses for hospitalization and death amongst HCWs who test positive and estimate the hazards according to vaccination status at the date of positive test, adjusting for covariates described in the case-control analyses. Sensitivity analyses will be conducted to evaluate the association of influence of a positive RT-PCR, antigen or serological test result prior to the vaccination campaign.

Sample size calculations and timing of analyses: The power of a matched case-control study depends on the assumed odds ratio and the number of discordant pairs (i.e. pairs in which the case is exposed and the control is unexposed, or vice versa), which is a function of the assumed odds ratio and the expected prevalence of exposure among controls. Moreover, the estimate of the odds ratio for one level of a categorical variable compared to baseline is determined by the distribution of all discordant pairs. As vaccine coverage and incidence are changing over time, the latter in ways we cannot predict, and there is no power formula for this analysis, we will simulate power and enroll individuals until we have reached a target power, which we can assess without analyzing the data. In particular, after determining the number of discordant case-control pairs for each combination of exposure categories, we will randomly assign one of each pair to each relevant exposure type according to a Bernoulli distribution, with the probability determined by the assumed odds ratio comparing the two categories. We will run an unadjusted conditional logistic regression on the simulated dataset to determine the p-value, and estimate the power as the proportion of N=1,000 simulations that return $p<0.05$. Code to perform the power calculation can be found at https://github.com/mhitchings/VEBRA_COVID-19.

Timing of interim and final analyses: Interim analyses will be performed to allow for early reporting of significant results for the benefit of public health, specifically the question whether receiving at least one dose of the vaccine is effective. For the primary outcome (symptomatic SARS-CoV-2 infection), we will perform an interim analysis of the effectiveness following at least one dose of the vaccine, as we expect this analysis to be the first to reach desired power. This interim analysis will be triggered upon reaching simulated power of 70% to detect vaccine effectiveness of 60% of at least one dose ≥14 days after the first dose. Once the interim analysis above has been triggered, we will perform one additional analysis when 80% power is achieved. To correct for multiple testing we will use the O'Brian Fleming alpha-spending method, meaning that for two interim analyses, the critical p-values at each analysis will be 0.0054 and 0.0492. We will perform final analyses of the primary outcome upon reaching simulated 80% power to detect vaccine effectiveness of 70% ≥14 days after the second dose.

Privacy: Only SEMSA, SES-AM and OPAS technicians had access to the identified dataset to linkage the datasets by name, date of birth, mother's name and CPF. After the linkage, the CPF was encrypted and the de-identified dataset was sent to the team for analysis.

Working group: Matt Hitchings, Otavio T. Ranzani, Julio Croda, Albert I. Ko, Derek Adam Cummings, Silvano Barbosa de Oliveira, Wildo Navegantes de Araujo, Jason R. Andrews, Ryan Borg, Roberto Dias de Oliveira, Daniel Barros, Andre Siqueira, Patricia Vieira da Silva, Bernardino Cláudio de Albuquerque, Shadia Hussami Hauache Fraxe, Cristiano Fernandes da Costa, Mario Sergio Sacaramuzzini Torres, Felipe Gomes Naveca, Vanderson de Souza Sampaio, Wade Schulz

**Supplementary Figure 1. Reported confirmed COVID-19 cases (Panel A) and COVID-19 associated deaths (Panel B) for case and death counts in the general population of the city of Manaus, Brazil** from March 2020 to April 1, 2021. Lines depict moving seven day averages for case and death counts**.** Data was obtained from the State Secretary of Health of Amazonas and collected by brasil.io. (ref).

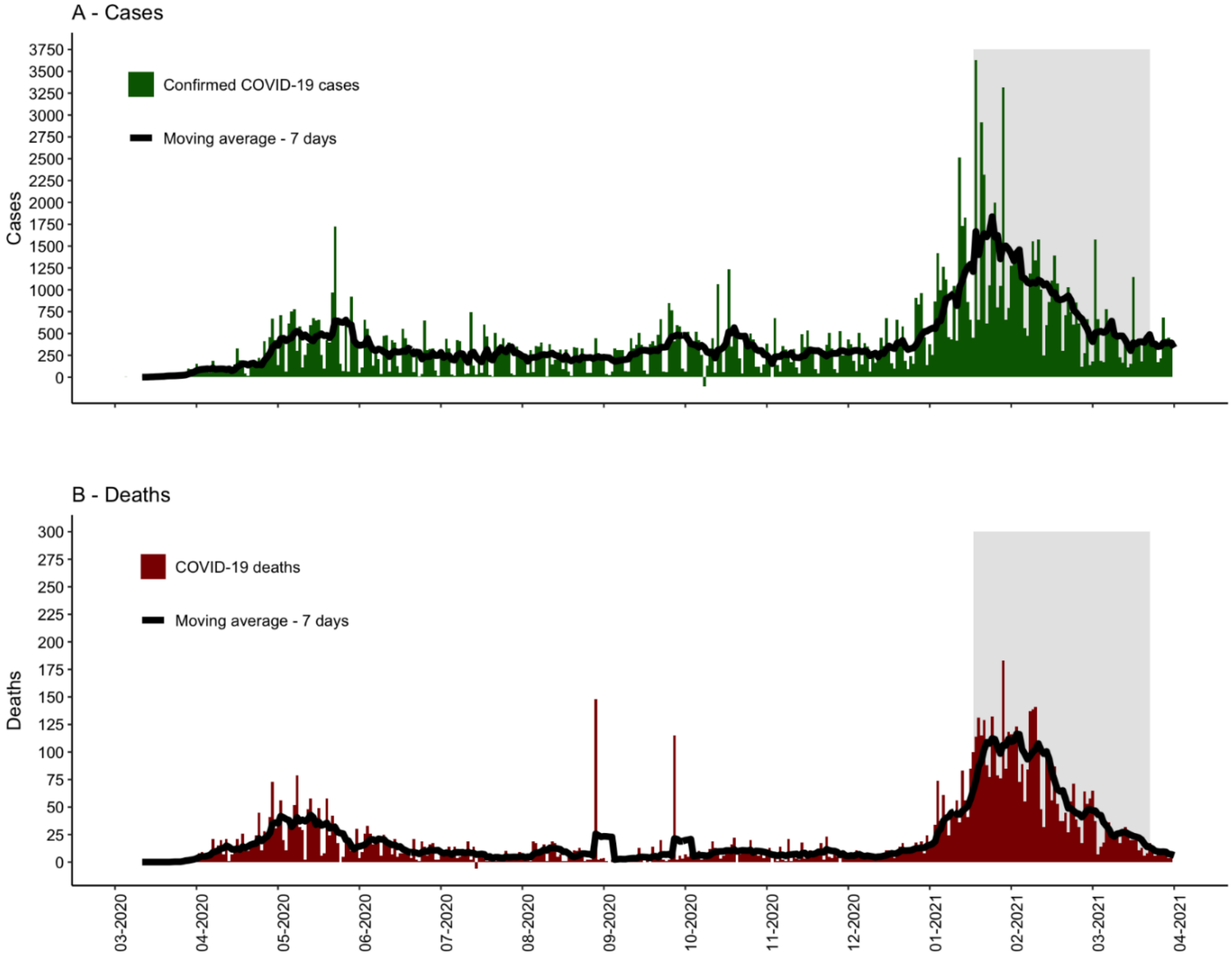

**Supplementary Figure 2: Proportion of P.1 variant among genotyped sequences in Manaus, from Nov 1 2020 to present.**

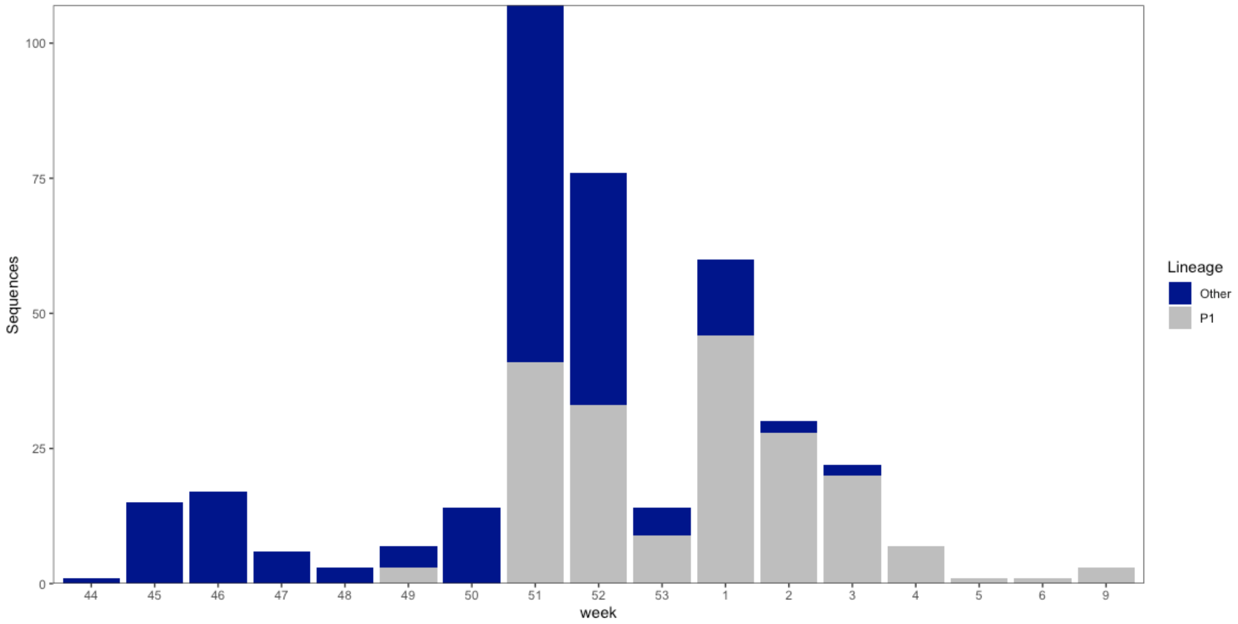

**Supplementary Figure 3.** PCR testing rate during the study period, PCR positive testing rate during the study period, test-positive proportion during the study period, and vaccine coverage, by age (from data extracted on April 14, 2021)

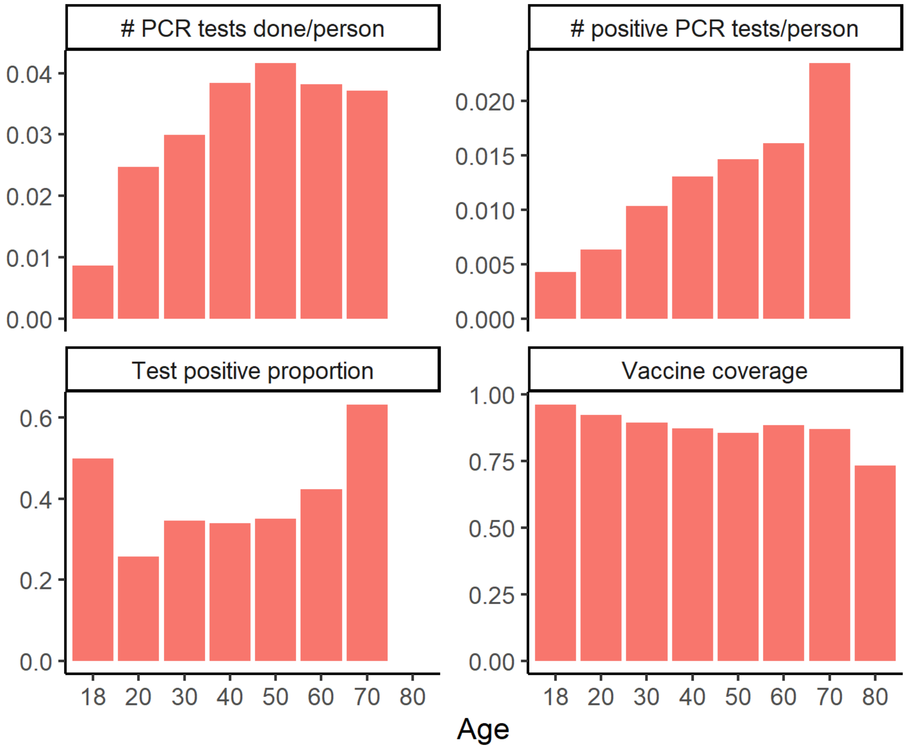

**Supplementary Figure 4.** PCR testing rate during the study period, PCR positive testing rate during the study period, test-positive proportion during the study period, and vaccine coverage by sex (from data extracted on April 14, 2021)

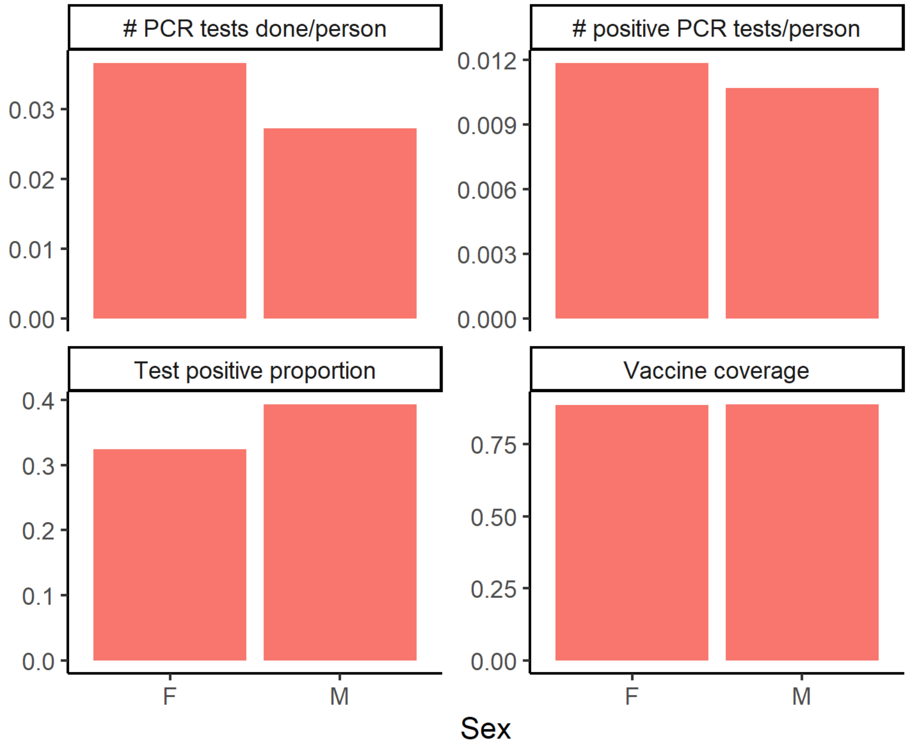

**Supplementary Figure 5.** PCR testing rate (left column) and PCR positive testing rate (right column) against average time from start of campaign to first dose administration, by neighbourhood (from data extracted on April 14, 2021). Top row represents PCR tests performed within the study period, and bottom row represents PCR tests performed since the start of the pandemic. Each point represents a neighbourhood. The average time to first dose administration is calculated from the start of the vaccination campaign, 19 January 2021.

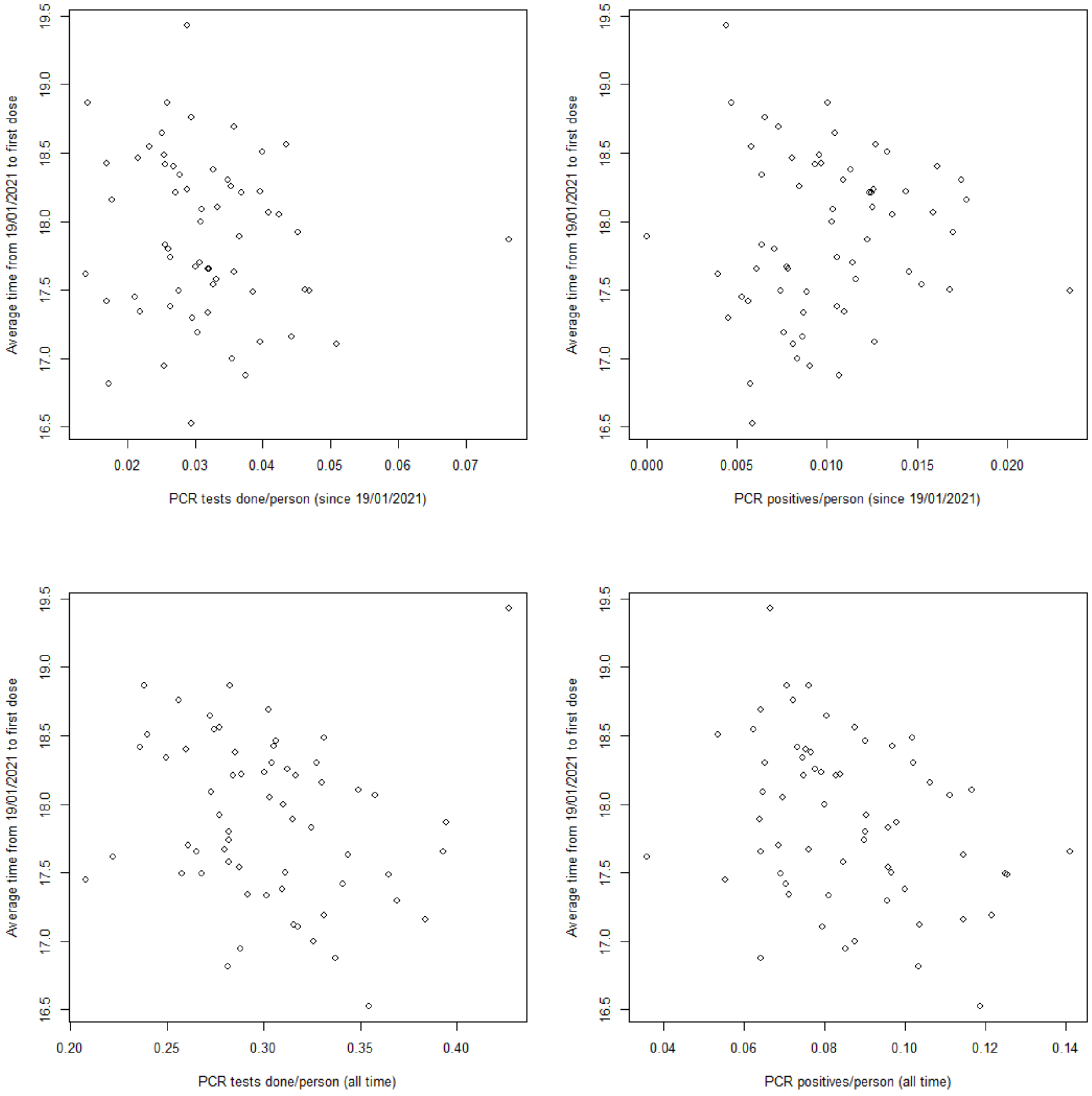

**Supplementary Figure 6.** Number of positive and negative PCR tests over time in the eligible study population, including selected case-control pairs and PCR tests not selected (from data extracted on May 4, 2021)

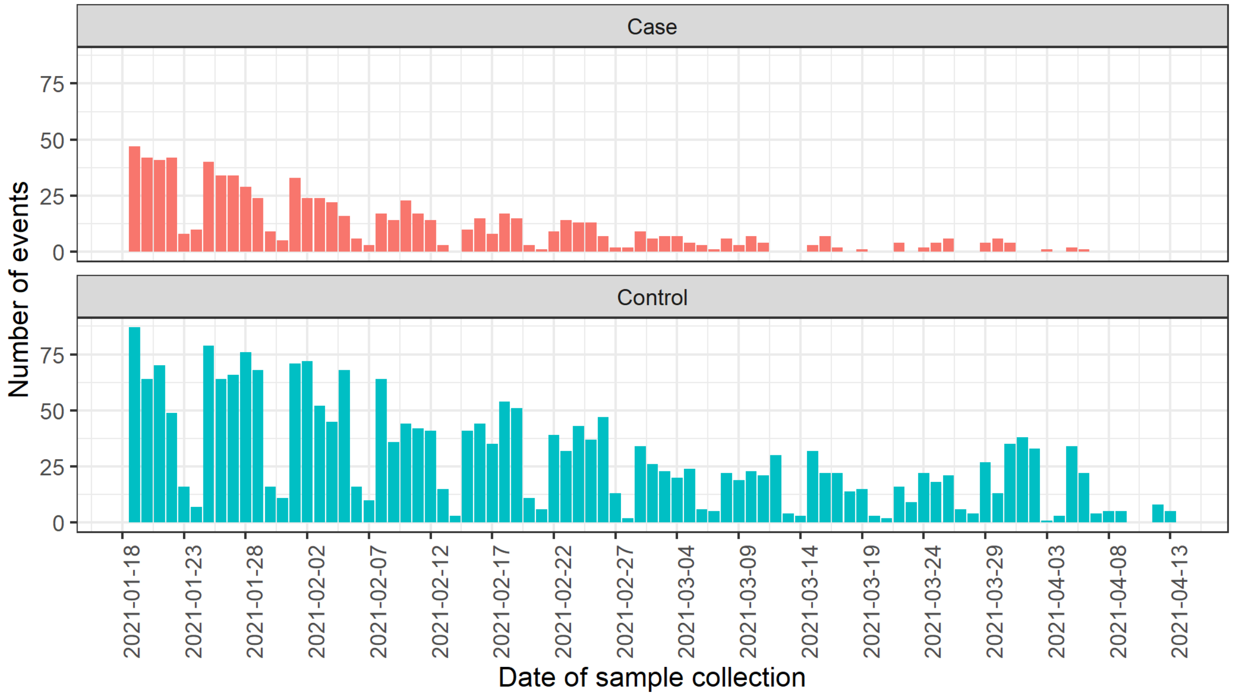

**Supplementary Figure 7: Timing of enrollment of discordant case-control pairs by vaccination category**

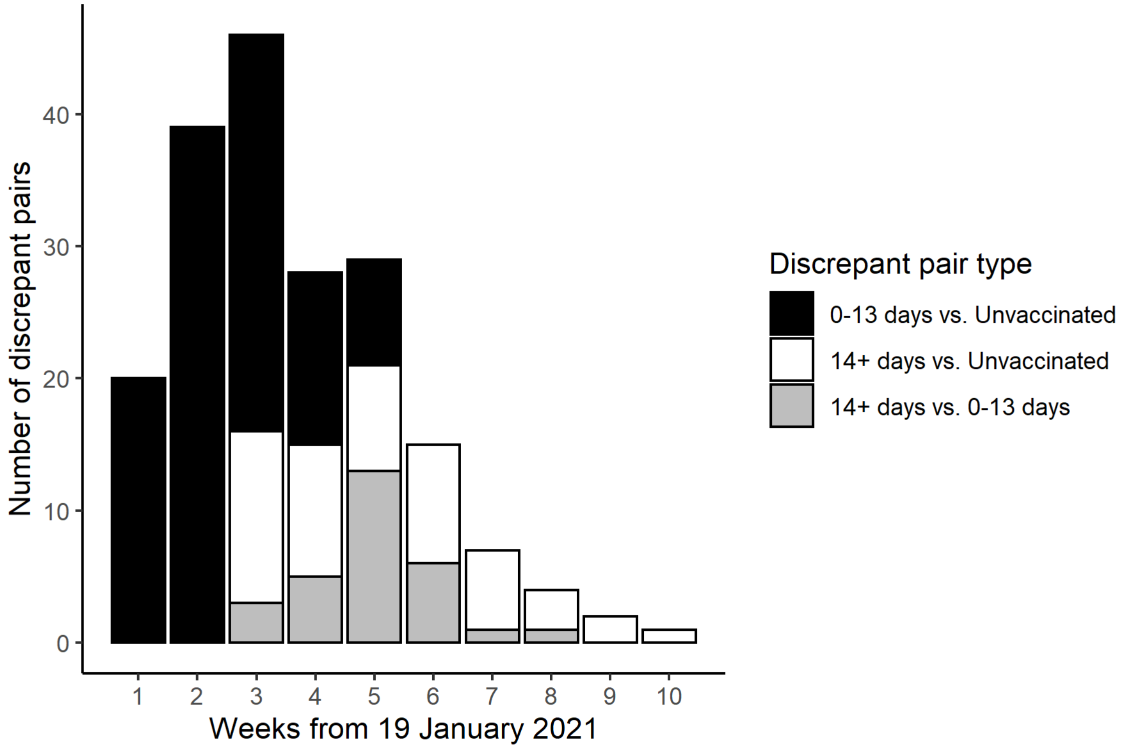

**Supplementary Figure 8: Timing of enrollment of discordant case-control pairs by vaccination category (two-dose analysis)**

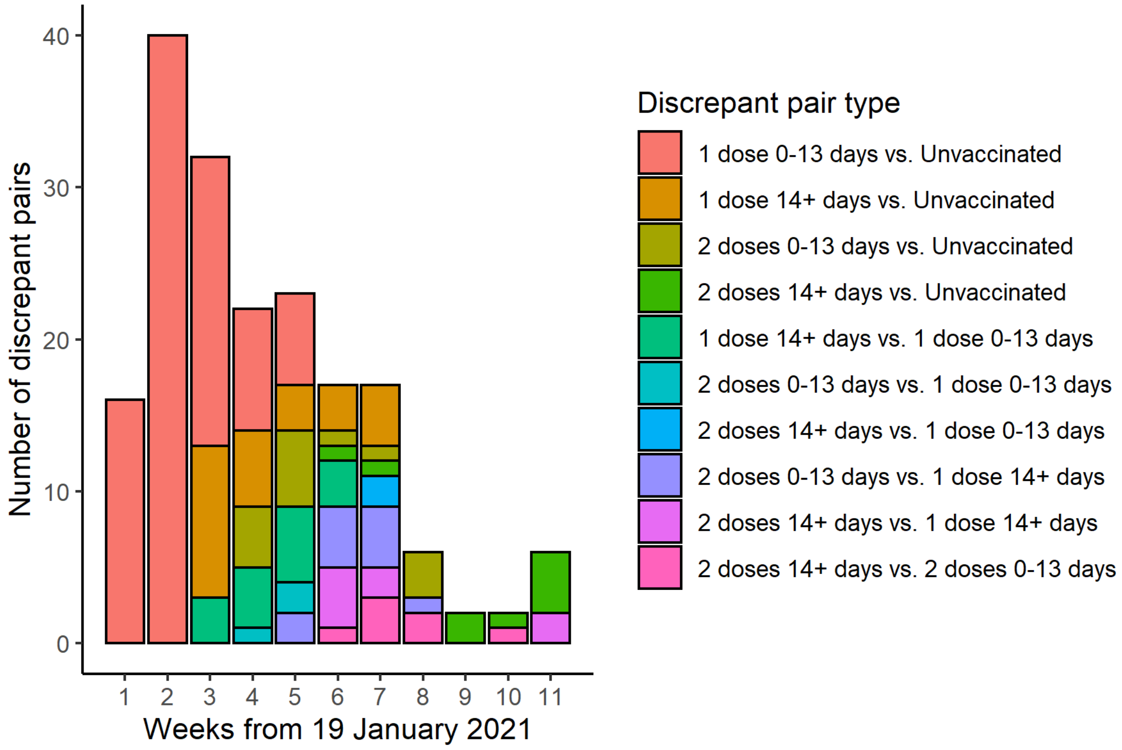

**Supplementary Figure 9: Timing of RT-PCR sample collection date relative to 1st (left column) and 2nd (right column) vaccine dose date, among cases (top row) and controls (bottom row) in the two-dose analysis, among individuals who were vaccinated during the study period**

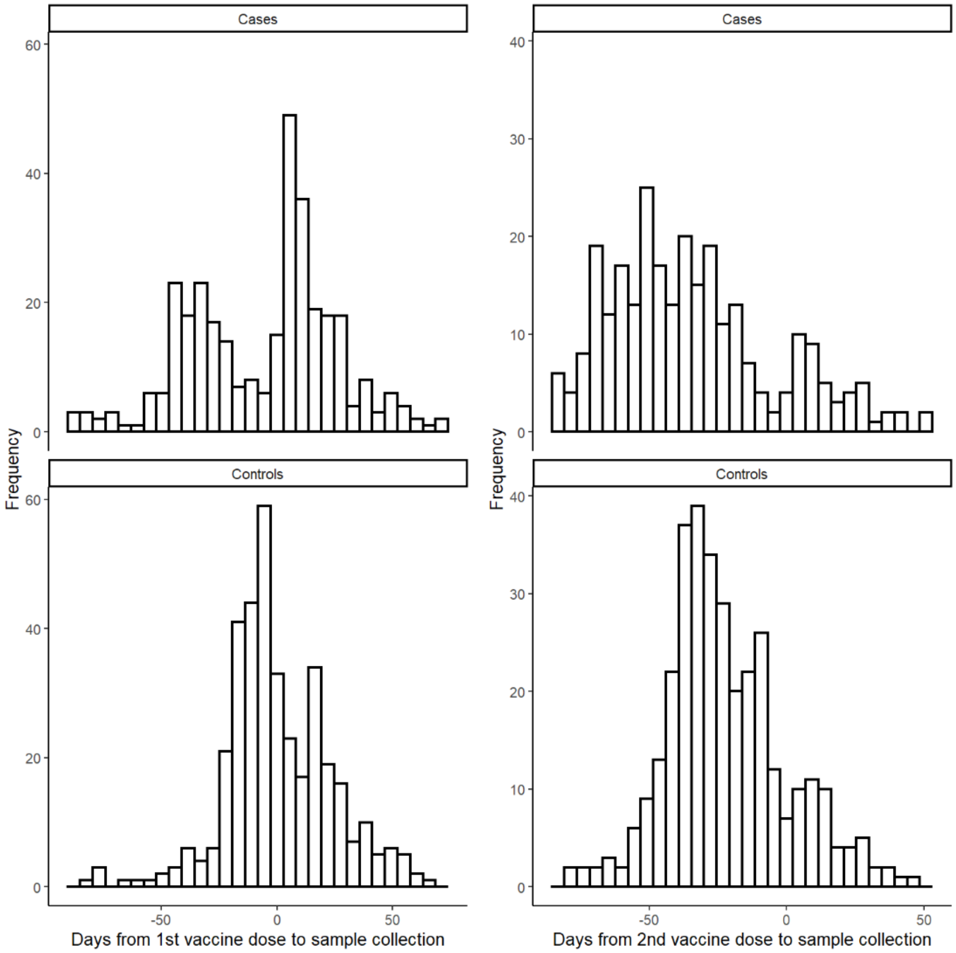

**Supplementary Table 1: Characteristics of matched vs. unmatched HCWs.**

| Characteristics | Matched (n=1,097) | Not matched (n=2,771) | Not eligible for case/control selection (n=63,850) |
|---|---|---|---|
| Vaccination | | | |
| Not vaccinated | 182 (17%) | 316 (11%) | 10,829 (17%) |
| Ever vaccinated | 915 (83%) | 2,455 (89%) | 53,021 (83%) |
| Age (years, mean (SD)) | 43.1 (9.3) | 41.5 (11.9) | 40.6 (11.9) |
| Female sex | 824 (74%) | 2,123 (77%) | 43,045 (67%) |
| Self-reported race/skin colour* | | | |
| Amarela/Yellow | 182 (16%) | 428 (15%) | 10,889 (17%) |
| Preta/Black | 14 (1%) | 31 (1%) | 710 (1%) |
| Pardo/Brown | 663 (60%) | 1,689 (61%) | 32,201 (50%) |
| Branca/White | 189 (17%) | 370 (13%) | 9,036 (14%) |
| Missing | 64 (6%) | 246 (9%) | 10,738 (17%) |
| Occupation category | | | |
| Administrative | 290 (26%) | 685 (25%) | 14,800 (23%) |
| Clinician | 36 (3%) | 120 (4%) | 3,373 (5%) |
| Nurse/nurse technician | 435 (39%) | 1,090 (39%) | 16,867 (26%) |
| Other health professional | 130 (12%) | 327 (12%) | 9,463 (15%) |
| Other health associated | 183 (17%) | 332 (12%) | 8,696 (14%) |
| Missing | 38 (3%) | 217 (8%) | 10,651 (17%) |
| Number of healthcare encounters[†] | 0.84 (1.1) | 0.73 (0.9) | 0.69 (0.9) |
| Prior positive SARS-CoV-2 test[†‡] | 53 (5%) | 257 (9%) | 4,363 (7%) |

**Supplementary Table 2: Case-control pairs for early analysis**

| | Case unvaccinated | Case ≥1 dose, 0-13 days | Case ≥1 dose, ≥14 days |
|---|---|---|---|
| Control unvaccinated | 172[1] | 51 | 15 |
| Control ≥1 dose, 0-13 days | 31 | 22 | 6 |
| Control ≥1 dose, ≥14 days | 28 | 16 | 52 |

[1] Concordant case-control pairs lie on the diagonal, with discordant pairs off the diagonal

**Supplementary Table 3: Discordant case-control pairs for two-dose analysis**

| | Case unvaccinated | Case 1 dose, 0-13 days | Case 1 dose, ≥14 days | Case 2 doses, 0-13 days | Case 2 doses, ≥14 days |
|---|---|---|---|---|---|
| Control unvaccinated | 178[1] | 58 | 9 | 5 | 5 |
| Control 1 dose, 0-13 days | 31 | 22 | 4 | 1 | 0 |
| Control 1 dose, ≥14 days | 16 | 11 | 17 | 5 | 3 |
| Control 2 doses, 0-13 days | 9 | 2 | 6 | 6 | 3 |
| Control 2 doses, ≥14 days | 4 | 2 | 5 | 4 | 12 |

[1] Concordant case-control pairs lie on the diagonal, with discordant pairs off the diagonal

**Supplementary Table 4:** Comparison of cases and controls without symptomatic illness

| | Early analysis | | Two-dose analysis | |
|---|---|---|---|---|
| **Characteristics** | **Cases (n=135)** | **Controls (n=135)** | **Cases (n=138)** | **Controls (n=138)** |
| Vaccination | | | | |
| Not vaccinated | 77 (57%) | 92 (68%) | 78 (57%) | 93 (67%) |
| One dose (first dose 0-13 days previously) | 32 (24%) | 17 (13%) | 32 (23%) | 17 (12%) |
| One dose (first dose ≥14 days previously) | 26 (19%) | 26 (19%) | 18 (13%) | 11 (8%) |
| Two doses (second dose 0-13 days previously) | | | 7 (5%) | 11 (8%) |
| Two doses (second dose ≥14 days previously) | | | 3 (2%) | 6 (4%) |
| Age (years, mean (SD)) | 43.6 (8.4) | 43.4 (9.0) | 43.7 (8.4) | 43.4 (9.0) |
| Female sex | 100 (74%) | 96 (71%) | 103 (75%) | 98 (71%) |
| Self-reported race/skin colour* | | | | |
| Amarela/Yellow | 20 (15%) | 25 (19%) | 21 (15%) | 25 (18%) |
| Preta/Black | 2 (2%) | 4 (3%) | 2 (1%) | 4 (3%) |
| Pardo/Brown | 85 (63%) | 79 (59%) | 87 (63%) | 82 (59%) |
| Branca/White | 14 (10%) | 16 (12%) | 14 (10%) | 16 (12%) |
| Missing | 14 (10%) | 11 (8%) | 14 (10%) | 11 (8%) |
| Occupation category | | | | |
| Administrative | 43 (32%) | 39 (29%) | 44 (32%) | 39 (28%) |
| Clinician | 3 (2%) | 6 (4%) | 3 (2%) | 6 (4%) |
| Nurse/nurse technician | 51 (38%) | 38 (28%) | 52 (38%) | 40 (29%) |
| Other health professional | 19 (14%) | 20 (15%) | 19 (14%) | 21 (15%) |
| Other health associated | 13 (10%) | 23 (17%) | 14 (10%) | 23 (17%) |
| Missing | 6 (4%) | 9 (7%) | 6 (4%) | 9 (7%) |
| Number of healthcare encounters† | 0.76 (1.05) | 1.09 (1.14) | 0.76 (1.04) | 1.10 (1.14) |
| Prior positive SARS-CoV-2 test†‡ | 1 (1%) | 17 (13%) | 1 (1%) | 19 (14%) |

*Race/skin colour as defined by the Brazilian national census bureau (Instituto Nacional de Geografia e Estatísticas) https://biblioteca.ibge.gov.br/visualizacao/livros/liv63405.pdf

†Prior to the start of the study on 19 January, 2021.

‡ Defined as SARS-CoV-2 RT-PCR or antigen detection.

**Supplementary Table 5: Vaccine effectiveness against all SARS-CoV-2 infection**

| | **Early analysis: All SARS-CoV-2 infection** | | **Primary analysis: All SARS-CoV-2 infection** | |
|---|---|---|---|---|
| | **OR (95% CI)** | **p-value** | **OR (95% CI)** | **p-value** |
| Unadjusted Analysis | | | | |
| 1 dose: 0-13 days after 1st vaccine dose vs. unvaccinated* | 1.81 (1.26-2.61) | <0.001 | 2.05 (1.43-2.94) | <0.001 |
| 1 dose: ≥14 days after 1st vaccine dose vs. unvaccinated* | | | 0.90 (0.54-1.50) | 0.68 |
| 2 doses: 0-13 days after 2nd vaccine dose vs. unvaccinated* | 0.68 (0.43-1.09) | 0.11 | 0.61 (0.31-1.20) | 0.15 |
| 2 doses: ≥14 days after 2nd vaccine dose vs. unvaccinated* | | | 0.54 (0.24-1.21) | 0.13 |
| Adjusted analysis | | | | |
| 0-13 days after 1st vaccine dose vs. unvaccinated* | 1.85 (1.26-2.71) | <0.001 | 2.16 (1.47-3.17) | <0.001 |
| ≥14 days after 1st vaccine dose vs. unvaccinated* | | | 0.88 (0.52-1.50) | 0.64 |
| 2 doses: 0-13 days after 2nd vaccine dose vs. unvaccinated* | 0.65 (0.40-1.07) | 0.09 | 0.50 (0.24-1.02) | 0.06 |
| 2 doses: ≥14 days after 2nd vaccine dose vs. unvaccinated* | | | 0.62 (0.26-1.46) | 0.28 |
| Age | 1.01 (0.99-1.03) | 0.24 | 1.01 (0.99-1.03) | 0.22 |
| Female sex | 0.69 (0.50-0.94) | 0.02 | 0.68 (0.50-0.92) | 0.01 |
| Self-reported race/skin colour[†] | | | | |
| Amarela vs. Pardo | 0.78 (0.54-1.14) | 0.21 | 0.81 (0.57-1.17) | 0.26 |
| Preta vs. Pardo | 1.06 (0.33-3.37) | 0.93 | 1.17 (0.36-3.8) | 0.79 |
| Branca vs. Pardo | 1.11 (0.78-1.59) | 0.56 | 1.03 (0.72-1.47) | 0.86 |
| Occupation category | | | | |
| ADM vs. Nurse | 1.03 (0.74-1.44) | 0.86 | 1.07 (0.77-1.49) | 0.68 |
| Clinical vs. Nurse | 1.44 (0.63-3.25) | 0.39 | 1.42 (0.63-3.17) | 0.39 |
| Other Health Associated vs. Nurse | 0.78 (0.52-1.17) | 0.22 | 0.79 (0.53-1.17) | 0.24 |
| Other Health Professional vs. Nurse | 0.79 (0.50-1.25) | 0.31 | 0.79 (0.51-1.23) | 0.30 |
| Prior healthcare encounters[‡] | | | | |
| 1-3 vs. 0 | 0.85 (0.65-1.11) | 0.23 | 0.9 (0.69-1.17) | 0.42 |
| ≥4 vs. 0 | 1.21 (0.54-2.69) | 0.65 | 1.91 (0.80-4.58) | 0.15 |
| Prior positive SARS-CoV-2 test[‡]** | 0.27 (0.13-0.55) | <0.001 | 0.26 (0.13-0.51) | <0.001 |

*At date of index sample collection for cases and controls

[†]Race/skin color is defined by IBGE (Instituto Nacional de Geografia e Estatísticas) https://biblioteca.ibge.gov.br/visualizacao/livros/liv63405.pdf

‡From March 2020 to 18 January, 2021
**Defined as SARS-CoV-2 RT-PCR or antigen detection

**Supplementary Table 6: Vaccine effectiveness asymptomatic SARS-CoV-2 infection**

| | Asymptomatic SARS-CoV-2 infection | |
|---|---|---|
| | **OR (95% CI)** | **p-value** |
| Unadjusted Analysis | | |
| 1 dose: 0-13 days after 1st vaccine dose vs. unvaccinated* | 2.82 (1.28-6.19) | 0.01 |
| 1 dose: ≥14 days after 1st vaccine dose vs. unvaccinated* | 2.41 (0.74-7.86) | 0.15 |
| 2 doses: 0-13 days after 2nd vaccine dose vs. unvaccinated* | 0.63 (0.11-3.78) | 0.61 |
| 2 doses: ≥14 days after 2nd vaccine dose vs. unvaccinated* | 0 (CI undefined) | - |
| Adjusted analysis | | |
| 0-13 days after 1st vaccine dose vs. unvaccinated* | 3.7 (1.48-9.22) | <0.001 |
| ≥14 days after 1st vaccine dose vs. unvaccinated* | 3.31 (0.92-11.94) | 0.07 |
| 2 doses: 0-13 days after 2nd vaccine dose vs. unvaccinated* | 0.65 (0.08-5.52) | 0.69 |
| 2 doses: ≥14 days after 2nd vaccine dose vs. unvaccinated* | 0 (CI undefined) | - |
| Female sex | 1.32 (0.74-2.38) | 0.35 |
| Prior positive SARS-CoV-2 test‡** | 0.04 (0-0.31) | <0.001 |

*At date of index sample collection for cases and controls

‡From March 2020 to 18 January, 2021

**Defined as SARS-CoV-2 RT-PCR or antigen detection

**Supplementary Table 7: Sensitivity analysis for vaccine effectiveness including comorbidities.** Vaccine effectiveness against symptomatic SARS-CoV-2 infection, and against all SARS-CoV-2 infection, with at least one dose and at least 14 days after administration of the first dose, including presence of one or more comorbidities as a covariate

| | **Primary outcome: Symptomatic SARS-CoV-2 infection** | | **Secondary outcome: All SARS-CoV-2 infection** | |
|---|---|---|---|---|
| | **OR (95% CI)** | **p-value** | **OR (95% CI)** | **p-value** |
| Unadjusted Analysis | | | | |
| 0-13 days after 1st vaccine dose vs. unvaccinated* | 1.61 (1.07-2.44) | 0.02 | 1.81 (1.26-2.61) | <0.001 |
| ≥14 days after 1st vaccine dose vs. unvaccinated* | 0.56 (0.32-0.95) | 0.03 | 0.68 (0.43-1.09) | 0.11 |
| Adjusted analysis | | | | |
| 0-13 days after 1st vaccine dose vs. unvaccinated* | 1.73 (1.11-2.7) | 0.02 | 1.87 (1.27-2.75) | <0.001 |
| ≥14 days after 1st vaccine dose vs. unvaccinated* | 0.52 (0.29-0.92) | 0.02 | 0.66 (0.40-1.08) | 0.1 |
| Age | 1.00 (0.98-1.03) | 0.64 | 1.01 (0.99-1.02) | 0.55 |
| Female sex | 0.53 (0.36-0.78) | <0.001 | 0.68 (0.5-0.93) | 0.02 |
| Self-reported race/skin color† | | | | |
| Amarela vs. Pardo | 0.84 (0.53-1.32) | 0.45 | 0.81 (0.55-1.18) | 0.27 |
| Preta vs. Pardo | 1.85 (0.39-8.75) | 0.44 | 0.98 (0.31-3.12) | 0.97 |
| Branca vs. Pardo | 1.22 (0.82-1.81) | 0.33 | 1.12 (0.79-1.61) | 0.52 |
| Occupation category | | | | |
| ADM vs. Nurse | 1.11 (0.74-1.64) | 0.62 | 1.04 (0.75-1.45) | 0.81 |
| Clinical vs. Nurse | 1.93 (0.72-5.17) | 0.19 | 1.35 (0.59-3.08) | 0.48 |
| Other Health Associated vs. Nurse | 0.96 (0.59-1.56) | 0.88 | 0.81 (0.53-1.23) | 0.32 |
| Other Health Professional vs. Nurse | 0.93 (0.55-1.58) | 0.79 | 0.80 (0.51-1.26) | 0.34 |
| Prior healthcare encounters‡ | | | | |
| 1-3 vs. 0 | 0.93 (0.67-1.29) | 0.66 | 0.78 (0.58-1.03) | 0.08 |
| ≥4 vs. 0 | 1.31 (0.51-3.37) | 0.57 | 1.10 (0.49-2.47) | 0.81 |
| Prior positive SARS-CoV-2 test‡ ** | 0.38 (0.17-0.87) | 0.02 | 0.26 (0.13-0.54) | <0.001 |
| Reported comorbidities | | | | |
| Any vs. none | 1.44 (0.95-2.18) | 0.09 | 1.36 (0.95-1.95) | 0.09 |

*At date of index sample collection for cases and controls

†Race/skin color is defined by IBGE (Instituto Nacional de Geografia e Estatísticas) https://biblioteca.ibge.gov.br/visualizacao/livros/liv63405.pdf

‡From March 2020 to 18 January, 2021

**Defined as SARS-CoV-2 RT-PCR or antigen detection
